# How to decide which COVID-19 patient with myocardial infarction to send to the Cath Lab? - A case series of COVID-19 patients with myocardial infarction

**DOI:** 10.1101/2021.02.07.21251081

**Authors:** Iana Simova, Denis Nikolov, Nikolay Dimitrov, Vladimir Kornovski, Vesela Tomova, Toni Vekov, Yordan Krasnaliev

## Abstract

**INTRODUCTION:** The Coronavirus pandemic has hit the world with its vast contagiousness, high morbidity, and mortality. Apart from the direct damage to the lung tissue, the corona virus infection is able to predispose patients to thrombotic disease, thus causing cerebral or coronary incidents.

**AIMS:** The aim of this study was to find a clinical or laboratory parameter, that would help in distinguishing between COVID-19 patients with myocardial infarction (MI), who have an infarct-related artery (IRA) and therefore, require immediate revascularization, and those, who have no IRA.

**METHODS:** This was a single-center, observational study of 10 consecutive patients with COVID-19, who were admitted with confirmed MI.

**RESULTS:** In our study group the mean age was 67.5 ± 8.3 years, half of the patients were female; all of them had arterial hypertension; 8 patients (80%) had dyslipidemy; 4 (40%) had diabetes. 30% of the patients with MI did not have an IRA, and did not require pPCI. Patients with MI and IRA had significantly higher hsTrI values (48.9 ± 43.2 vs 0.6 ± 0.7, p=0.007) and exclusively typical chest pain 100% vs 0%, p=0.007), compared to patients with MI without an IRA. The ECG changes had only marginal statistical significance. Our results suggest that using a higher cut-off value for hsTrI (>7.5 times upper reference range) increases the specificity and positive predictive value for diagnosing a MI with the presence of IRA and need for pPCI, to 100%

**CONCLUSION:** In our analysis we confirm that a higher cut-off value for hsTrI helps distinguish between COVID patients with MI, who have IRA and therefore, require immediate revascularization, compared to those, who have no IRA.

## INTRODUCTION

The coronavirus pandemic has hit the world with its vast contagiousness, high morbidity, and mortality **[1]**. Apart from the direct damage to the lung tissue, the corona virus infection is associated with multiple organ damage, including the heart. Emerging evidence reveals a direct correlation between COVID-19 and cardiovascular complications, such as heart failure, myocarditis, arrhythmias, conduction abnormalities and acute coronary syndromes **[2]**. The SARS-CoV-2 infection can frequently induce coagulation abnormalities that are associated with cardiopulmonary deterioration and death as a possible complication in all patients, despite presence or absence of concomitant risk factors and diseases. In addition, many patients with severe COVID-19 undergo thromboembolic events, which seem to be related to this particular coagulopathy **[3][4]**. One of the most unpleasant and life-threatening types of thromboembolism is the one involving the coronary circulation, thus causing a heart attack. Many additional problems arise due to this condition e.g., access to a Cath lab, exposure of additional medical personnel, more complications and increased mortality for the patients.

Invasive angiography for COVID-19 patients is logistically challenging and, in some cases, there is no intervention target, since microcirculatory disease and thrombosis is common in this group. Therefore, we studied in detail the case series of 10 patients referred for primary percutaneous coronary intervention (pPCI) for MI in our catheterization laboratory during the course of COVID-19 infection **[5]**. And we set ourselves the purpose to evaluate if there are some factors or parameters that could predict the presence of an interventional target – infarct related artery (IRA), prior to catheterization, and to determine their sensitivity and specificity.

## MATERIALS AND METHODS

The COVID-19 department of Heart & Brain University Hospital, Pleven, Bulgaria, functions since 11.2020, with 64 beds, 24 of which are intensive with the option for mechanical ventilation. For the last two months of 2020, 214 patients were treated in our COVID-19 department. Ten of them were referred to the catheterization laboratory for selective coronary angiography with myocardial infarction (MI), defined by the third universal definition of MI **[6]**. Most of our patients were directed to our hospital with ACS as their diagnosis, while others developed ACS during their stay in the COVID department and were therefore brought to the Cath lab. During the procedure, appropriate personal protective equipment (PPE) is worn by the medical personnel, including a sterile gown, gloves, goggles and a N95 mask. The patient is brought to the Cath lab trough a different one-way corridor, in order to reduce chances of infection. The angiography includes a standard set of diagnostic and guiding catheters, mainly EBU 3.5/6Fr for the left coronary artery, and JR3.5 for the right, coronary guidewires, drug eluting stents and balloons. The majority of the catheterization laboratories have either normal or positive ventilation systems and are not designed to contain an infectious environment. Therefore, catheterization laboratories will require a thorough disinfection following every procedure, leading to delays for the scheduled procedures.

### Statistical analyses

Statistical analyses were performed using SPSS statistical software for Windows version 19.0. The distribution of continuous variables was tested using the Kolmogorov-Smirnov test. Normally distributed data were presented as mean ± standard deviation (SD), whereas non-normally distributed data – as median and interquartile range (IQR) (the difference between the 25th and 75th percentile). Categorical variables were presented in percentage terms. We compared differences between groups with Independent-Samples T-Test. Sensitivity, specificity, positive and negative predictive values (PPV and NPV) were calculated according to the true positive (TP), false positive (FP), true negative (TN) and false negative (FN) results, using the following formulas: Sensitivity = TP/(TP+FN); Specificity = TN/(TN+FP); PPV = TP/(TP+FP); NPV = TN/(TN+FN). A two-tailed p value < 0.05 was considered statistically significant.

### Ethics

All patients signed an informed consent for coronary angiography and PCI, and for personal data analysis. The study protocol is in accordance with the Declaration of Helsinki.

## RESULTS

Mean age in our group was 67.5 ± 8.3 years; half of the patients were female; all patients had arterial hypertension; 8 patients (80%) had dyslipidemy; 4 (40%) had diabetes. On admission all patients had chest pain [typical in 7 (70%), atypical in 2 (20%) and non-stenocardic in 1 (10%)] and an increase in serum level of high-sensitive Troponin I (hsTrI).

Five of the patients (50%) had ST segment elevation, typical for ST elevation myocardial infarction (STEMI); one person (10%) had a new onset left bundle branch block (LBBB); 1 (10%) had significant ST depression and 3 (30%) had no significant ECG changes.

Three of the patients (30%) had heart failure symptoms on admission. Before developing symptoms and signs of MI, most of the patients (6 – 60%) were hospitalized and treated for COVID-19 at our COVID-department, 2 (20%) were referred from other hospitals with COVID-departments, and 2 (20%) were referred from emergency care units after being treated for COVID-19 at home. The mean hospital stay before the acute coronary syndrome (ACS) was 11.3 ± 10.8 days; 2 patients were in Intensive care unit (ICU) and they were on mechanical ventilation. All but one person in our group were discharged alive.

After coronary angiography, we found that 7 patients (70%) had an infarct related artery / lesion (IRA) and they underwent pPCI. The other 3 (30%) did not have an IRA, pPCI was not performed, and the diagnosis of myocardial infarction with no obstructive coronary arteries (MINOCA) was made, most probably due to myocarditis.

Comparing the patients with IRA to those without we found that the subjects who finally required pPCI had significantly higher hsTRI values, exclusively had typical chest pain (7 out of 7 patients with IRA had typical chest pain, while the other three without IRA had an atypical or non-stenocardic chest pain, with significant difference between the groups regarding characteristics of chest pain: p=0.007), and had more often ST elevation (5 out of 7 patients with IRA had ST elevation, 1 had new LBB and 1 had no ECG changes, while 1 out of 3 patients without IRA had ST depression and 2 had no ECG changes, with a borderline significance between the groups regarding ST segment changes: p=0.05). The other studied variables did not differ significantly between the groups with or without IRA – table 1.

**Table 1.** Comparison between the groups with and without an IRA and need for a PCI

| Variable | Patients with IRA and pPCI<br>n = 7 | Patients w/o IRA and pPCI<br>n = 3 | p<br>value |
| --- | --- | --- | --- |
| Age (years) (mean ± SD) | 67.1 ± 9.2 | 68.3 ± 7.4 | 0.48 |
| Male (n, %) | 3 (43%) | 2 (66%) | 1.0 |
| AH (n, %) | 7 (100%) | 3 (100%) | 1.0 |
| DLP (n, %) | 5 (71%) | 3 (100%) | 1.0 |
| DM (n, %) | 2 (29%) | 2 (66%) | 0.5 |
| Typical chest pain (n, %) | 7 (100%) | 0 (0%) | 0.007 |
| ST elevation (n, %) | 5 (%) | 0 (0%) | 0.05 |
| Symptoms of HF (n, %) | 2 (29%) | 1 (33%) | 1.0 |
| Symptom onset (days) (mean ± SD) | 14.8 ± 8.7 | 12.3 ± 7.6 | 0.47 |
| Home treatment (n, %) | 2 (29%) | 1 (33%) | 0.87 |
| SatO<sub>2</sub> at admission (%) (mean ± SD) | 83.7 ± 10.7 | 85.5 ± 9.3 | 0.85 |
| Hospital stay (days) (mean ± SD) | 14 ± 13.3 | 4.5 ± 2.1 | 0.12 |
| ICU stay (days) (mean ± SD) | 9 ± 8.4 | 0 (0%) | 0.057 |
| Mechanical ventilation (n, %) | 2 (29%) | 0 (0%) | 1.0 |
| hsTnI (ng/ml) (mean ± SD) | 48.9 ± 43.2 | 0.6 ± 0.7 | 0.007 |
| CK (U/l) (mean ± SD) | 288.7 ± 341.1 | 177.7 ± 184 | 0.21 |
| CK-MB (U/l) (mean ± SD) | 43.9 ± 51.5 | 13.7 ± 5 | 0.13 |
| D-dimer ((ng/ml) (mean ± SD) | 3487.7 ± 6411.4 | 962.7 ± 835.9 | 0.24 |
| hsCRP (mg/l) (mean ± SD) | 81.6 ± 84.3 | 116.7 ± 75 | 0.55 |
| Leu ($\times 10^9$ g/l) (mean ± SD) | 12.9 ± 5.5 | 7.6 ± 3.4 | 0.52 |
| Lym ($\times 10^9$ g/l) (mean ± SD) | 1.1 ± 0.6 | 0.7 ± 0.4 | 0.26 |
| LDH (U/l) (mean ± SD) | 1015 ± 722.1 | 958.7 ± 512.5 | 0.58 |
| ASAT (U/l) (mean ± SD) | 144.1 ± 114.8 | 108 ± 128.3 | 0.99 |
| ALAT (U/l) (mean ± SD) | 105.1 ± 125.3 | 59.7 ± 68.9 | 0.66 |

Regarding hsTrI concentrations [the upper reference limit (URL) in our laboratory is 0.2 ng/ml], all but one patient with IRA and pPCI had hsTrI >1.5 ng/ml (>7.5 times URL), and all patients without IRA and pPCI had hsTrI ≤1.5 ng/ml (≤7.5 times URLN). Therefore, for hsTrI >1.5 ng/ml (>7.5 times URL) to predict the presence of IRA and the need for pPCI the sensitivity is 86%, the specificity is 100%, positive predictive value (PPV) is 100%, while the negative predictive value (NPV) is 10%.

## DISCUSSION

Myocardial infarction, defined by the third universal definition of MI, could complicate up to 5% of COVID-19 cases. In our study group, 30% of the patients with MI did not have an IRA and, consequently, did not need a coronary intervention. Patients with MI and IRA had significantly higher hsTrI values and exclusively typical chest pain, compared to patients with MI but without an IRA, whose hsTrI values were lower and chest pain was atypical or non-stenocardic. ECG changes had only a minor statistical significance for distinguishing between MI patients with or without IRA. Our results suggest that using a higher cut-off value for hsTrI increases the specificity for diagnosing a MI and therefore - interventional treatment.

According to the third universal definition of myocardial infarction the diagnosis requires evidence of myocardial necrosis in a clinical setting consistent with acute myocardial ischemia. These criteria require detection of a rise and/or fall in cardiac biomarker levels (preferably cardiac troponin) with at least one value above the 99th percentile upper reference limit, with at least one of the following: symptoms of myocardial ischemia, new or presumed new significant ST-segment T-wave changes or new left bundle branch block, development of pathological Q-waves on the ECG, imaging evidence of loss of viable myocardium or new regional wall motion abnormality or identification of intracoronary thrombus by angiography or autopsy.

This universal definition of MI, however, might not be the optimal guide to send a patient to the catheterization laboratory in the setting of procoagulation abnormalities in the course of acute or post-acute COVID-19.

The range of clinical responses to COVID-19 is extremely broad. Endothelial injury is an underlying mechanism that links the inflammation and consequent thrombosis **[7], [8]**. It is currently hypothesized that ACE-2 receptor is the entry gate for the virus to invade and infect tissues. The vascular endothelium appears to be targeted directly by the virus as ACE-2 is expressed widely in the blood vessels and the heart. The result is exocytosis of endothelial granules containing VWF (von Willebrand factor), P-selectin, and other proinflammatory cytokines, which mediate platelets adhesion, aggregation, and leukocyte adherence to the vessel wall, with a final result of intravascular thrombosis **[9]**.

Even though our analysis is on a small number of patients, similar incidence of arterial (coronary and cerebral) thrombosis (4%) has been described by other authors. In this study, however, the authors have not provided a guide to the right moment of interventional treatment. According to our published data search, we were not able to find another study, analyzing the predictors for the presence of IRA and the need for pPCI in COVID-19 MI patients.

## CONCLUSION

In our analysis we confirm that a higher cut-off value for hsTrI helps distinguish between COVID patients with ACS, who have IRA and therefore, require immediate revascularization, compared to those, who have no IRA.

## Data Availability

Statistical analyses were performed using SPSS statistical software for Windows version 19.0. The
distribution of continuous variables was tested using the Kolmogorov-Smirnov test. Normally distributed
data were presented as mean +/− standard deviation (SD), whereas non-normally distributed data - as
median and interquartile range (IQR) (the difference between the 25th and 75th percentile). Categorical
variables were presented in percentage terms. We compared differences between groups with
Independent-Samples T-Test. Sensitivity, specificity, positive and negative predictive values (PPV and
NPV) were calculated according to the true positive (TP), false positive (FP), true negative (TN) and false
negative (FN) results, using the following formulas: Sensitivity = TP/(TP+FN); Specificity = TN/(TN+FP);
PPV = TP/(TP+FP); NPV = TN/(TN+FN). A two-tailed p value < 0.05 was considered statistically significant.

